# Predictors of CPAP outcome in hospitalised COVID-19 patients

**DOI:** 10.1101/2020.06.14.20130880

**Authors:** Y Noeman-Ahmed, S Gokaraju, DJ Powrie, DA Amran, I El Sayed, Ashraf Roshdy

## Abstract

**Introduction:** Throughout March – April 2020, many patients with COVID-19 presented to Southend University Hospital with Acute Hypoxaemic Respiratory Failure (AHRF). Patients were managed in a Specialist Respiratory High Dependency Unit. We present our experience on the usage of continuous positive airway pressure (CPAP) therapy and possible indicators of its success in this patient group.

**Methods:** Data from patients (n=89) requiring mechanical ventilation during the months of March-April 2020, were retrospectively collected and analysed. 37 patients received IMV (Invasive Mechanical Ventilation) without a CPAP trial beforehand. 52 patients underwent a CPAP trial, of which 21 patients successfully avoided intubation and ITU admission.

**Results:** The 52 patients, prior to receiving CPAP had significant respiratory failure as evidenced by a low PaO2: FiO2 (PFR) (mean± SD 123 ± 60 mmHg) and mean SpO2:FiO2 (SFR) (mean ± SD: 140 ± 50). The main indicators of CPAP success were: higher SFR before and after CPAP, lower respiratory rate (RR), lower Neutrophil to Lymphocyte ratio (NLR) and higher PFR prior to CPAP.

**Discussion:** CPAP proved successful in 40% of COVID-19 patients presenting with AHRF. SFR, PFR, RR and NLR are predictors of such success. SFR can be used for effective real time monitoring of patients before and after CPAP to identify likelihood of success. Based on our results, we have suggested a modified CPAP management protocol in COVID-19. These findings can guide future studies and will allow improved triage of patients to either CPAP or IMV, in the event of a future COVID peak.

## Introduction

As of May 2020, more than 5.9 million people have been diagnosed with COVID-19 including more than 367,000 fatalities. **(1)** Whilst most patients are asymptomatic or mildly affected, in the United Kingdom, 17% of COVID-19 hospitalised patients were admitted to Intermediate or Intensive Care units. (2)

In COVID-19, acute hypoxaemic respiratory failure (AHRF) may be caused by acute respiratory distress syndrome (ARDS) or pneumonia (not meeting the ARDS criteria). (3) In cases refractory to oxygen therapy, the view that such patients should undergo early invasive mechanical ventilation (IMV) without the option for less invasive treatments such as non-invasive positive pressure ventilation NIPPV has been suggested. (4) However, IMV is not without complications, such as ventilator-induced lung injury (VILI), excessive sedation and haemodynamic consequences. (5) NIPPV can be provided as continuous positive airway pressure (CPAP) or bi-level positive airway pressure (BiPAP). (6) The role of CPAP in AHRF in COVID-19 is currently not well established. Currently in the UK, 16% and 10% of UK hospitalised COVID-19 patients have received IMV and NIPPV respectively. (2)

The UK National Health Service (NHS) issued specific guidance on the use of NIPPV in COVID-19 in March 2020. Following this guidance, we created an algorithm for the use of CPAP in COVID-19 patients admitted to our hospital (Figure 1). The decision of whether to give the patient a trial of CPAP or to intubate directly was guided by our algorithm. However, it is important to note that the final decision lay with the on call respiratory consultant and the ITU team. We conducted a retrospective cohort study to investigate the predictors of CPAP outcome in these patients.

**Figure (1):**
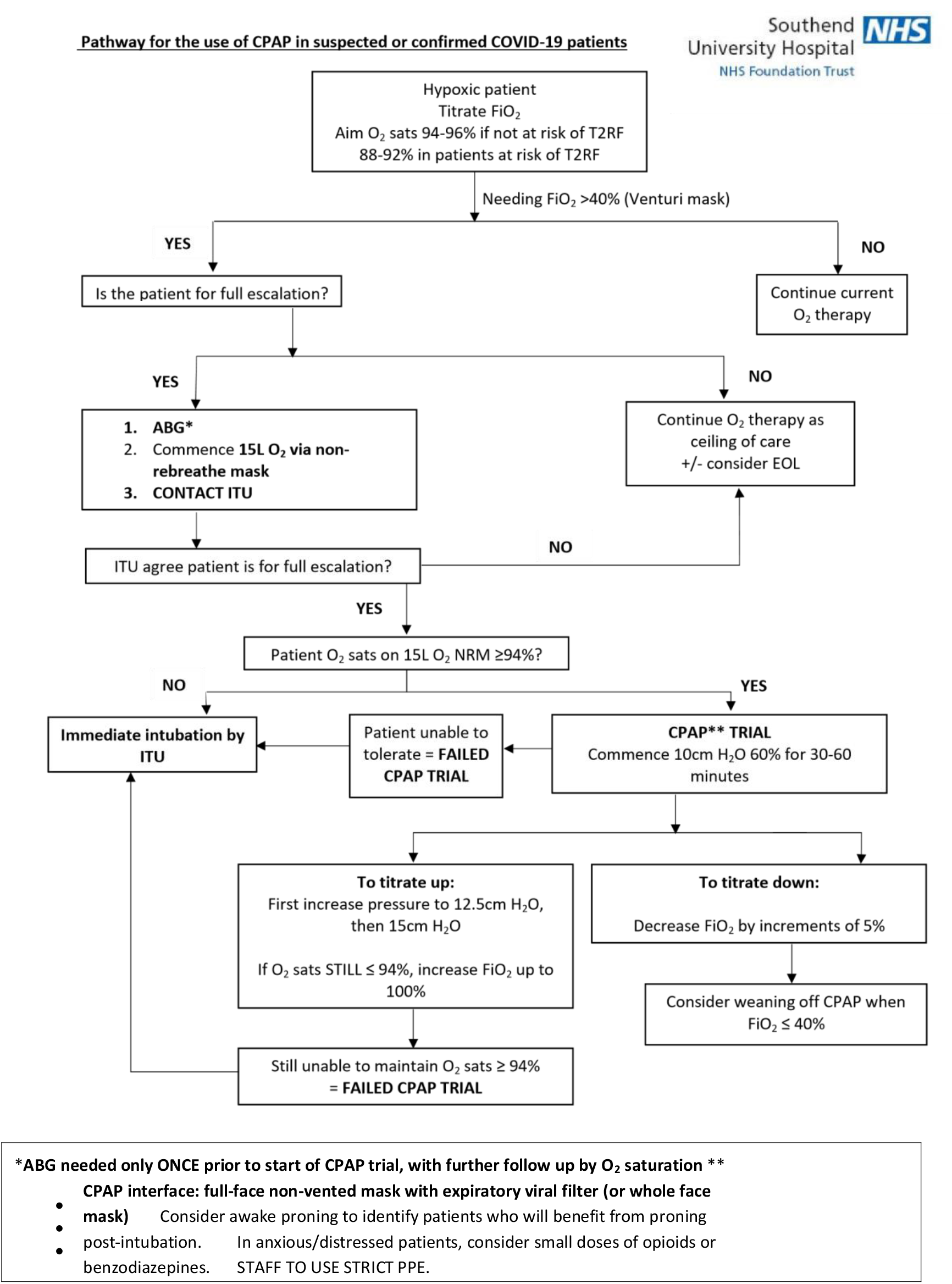
Pathway for the use of CPAP in suspected or confirmed COVID-19 patients. Contact CPAP Respiratory consultant on Rochford ward (ext 5474, 5475) if further advice needed

## Patients and methodology

### Inclusion Criteria

Adult COVID-19 patients (≥ 18 years old) admitted to Southend University Hospital (UK) between the 1^st^ of March 2020 and the 30^th^ of April 2020 that underwent CPAP treatment in the Acute Respiratory Care Unit (ARCU) were included in the study. All patients had been tested SARS-CoV2 RNA positive using Nose and Throat swabs sent for real time RT-PCR (RdRp gene assay).

### Exclusion criteria

COVID-19 patients who had any form of domiciliary NIPPV for obstructive sleep apnoea, or for any other reason, prior to hospital admission, were excluded from the study. Patients who underwent hospital BiPAP or received CPAP for the treatment of cardiogenic pulmonary oedema were also excluded from the study.

### Settings

The ARCU is a specialised unit staffed by experienced respiratory consultants and nurses. Patients on CPAP were nursed with 1:2 nurse to patient ratio. Emphasis was placed on a secure fitting interface of the CPAP system with a double filter (expiratory filter for the face mask, and exhaust system filter) in order to reduce widespread dispersion of exhaled air in addition to strictly following guidance on use of personal protective equipment (PPE), thus minimising the risk of airborne generating procedures (AGPs) to healthcare workers. (4)

### Data Collection

Data was collected retrospectively using various hospital databases and patients’ medical notes. The ARCU reports to the Intensive Care National Audit and Research Centre (ICNARC). We screened the ICNARC database for all eligible COVID-19 patients and collected their demographic data. Patients escalation status was collected using ‘treatment escalation plan’ prior to CPAP trial and patients were classed as ‘full escalation’ or ‘NIPPV’ ceiling of treatment.

### Vital signs & Investigations

The patients’ oxygen saturations (SpO_2_) on room air as recorded by pulse oximetry was recorded on admission. Patients oxygen saturations (SpO_2_) and supplementary oxygen (FiO_2_) prior to CPAP were recorded. Respiratory rate (RR), heart rate (HR) and alertness on AVPU scale were also recorded.

SpO_2_ to FiO_2_ ratio (SFR) was calculated prior to receiving CPAP. SFR post CPAP trial was also recorded using an average of the first three pulse oximeter recordings divided by the average of the first three FiO_2_ recordings within 30-120 minutes of the start of CPAP. All measurements were taken while patient was in the supine position, and when on CPAP, its pressure was 10 cm H2O.

The PFR (mmHg), also known as the Horowitz index was calculated as the ratio between PaO_2_ (in mmHg) and FiO_2_. In order to standardise FiO_2_ across various oxygen delivery systems, the following standardisations were used:

1. Prior to CPAP: Most of the patients were on Venturi Mask 40% as per protocol. Other oxygen delivery systems were calculated as follows: High flow oxygen via Whisperflow, Philips (100%), Non rebreathe mask 15 L/m flow (85%) and other standard venturi denominations 2 L/min (24%), 4 L/min (28%), 8 L/min (35%), 10L/min (40%) 15 L/min (60%)
2. On CPAP: FiO_2_ readings on the oxygen analyser of the Whisperflow Philips CPAP machine were recorded. For the minority of patients where CPAP mode of the NIPPY 3+ machine was used with supplemental oxygen, calculations followed the Venturi protocol, i.e. 10L (40%), 15L/min (60%). (7)

The following biochemical markers were recorded upon admission and prior to CPAP: CRP (mg/L), Neutrophils (10^9^/L), Lymphocytes (10^9^/L) and D-dimer (ng/ml). Neutrophil to lymphocyte ratio (NLR) was calculated. In addition, admission chest x-ray findings were reviewed retrospectively by a radiologist to determine if they showed bilateral pulmonary infiltrates, compatible with the diagnosis of ARDS.

### End point

A successful CPAP trial was defined as the successful weaning of CPAP to other oxygen delivery devices with no re-institution of any form of MV for 72 hours. A ‘fail’ on the CPAP trial occurred when the responsible respiratory consultant decided to end the CPAP trial, either proceeding to intubation or if their ceiling of treatment was NIPPV they were stepped down to a ward-based ceiling of treatment.

### Registration and ethical approval

The work has been approved ethically and registered in the hospital R&D under registration number 20007.

### Statistical Analysis

Quantitative data were described by mean (standard deviation) and median (minimum-maximum). While categorical variables were summarized by frequency and percent.

Receiver Operating Characteristic (ROC) curve analysis by DeLong method was performed to detect the predictive performance of different indices for CPAP success. (8) An optimal cut-off point for each index test was determined by Youden index. (9)

We conducted bivariate analysis using Independent sample t-test or Mann-Whitney test based on quantitative variables distribution as well as Pearson’s Chi-square test to compare different demographic and clinical parameters between CPAP success and failure groups. Statistically significant, and clinically relevant categorized predictors based on ROC analysis results were fitted in multivariate stepwise backward Wald logistic regression analysis. Pearson’s correlation test was also used to study linear relation between PFR and SFR. (10)

Decision trees by classification and regression tree (CART) analysis were performed for depicting the prognostic factors associated with success before and after CPAP. The main variables associated with CPAP success were ordered by their relative importance: SFR before and after CPAP, respiratory rate prior to CPAP, NLR and PFR. Performance of each classification tree was determined by calculating accuracy of model’s predicted probability with corresponding area under the curve. (11)

We did not calculate sample size, however, power analysis for each of AUC for SFR before and after CPAP as well as NLR was at least more than 80%.

Statistical analysis was done using IBM SPSS statistics program (12), and R software with the following packages: “rpart”, “rpart.plot”, “pROC” and “ROCR”. (13) All statistical tests were two-sided and judged at 0.05 significance level.

## Results

89 patients with AHRF were identified. 37 patients were intubated directly and 52 patients underwent a CPAP trial in the ARCU. 40.3% of the patients passed the CPAP trial (n=21). All patients survived after passing the CPAP trial while among those intubated 38% died (p<.001).

There were no significant differences in demographic data or co-morbidities between those groups who failed or succeeded on CPAP (Table 1). Diabetes Mellitus was more prevalent in the CPAP failure group (32% vs 14%) but that difference did not reach statistical significance. The CPAP cohort suffered significant respiratory failure as evidenced by a mean PFR (123.15 ± 59.56) and mean SFR (140 ± 50). 48 patients (92%) met the Berlin definition of ARDS confirming the severity of the disease in this group of patients.

**Table 1:**
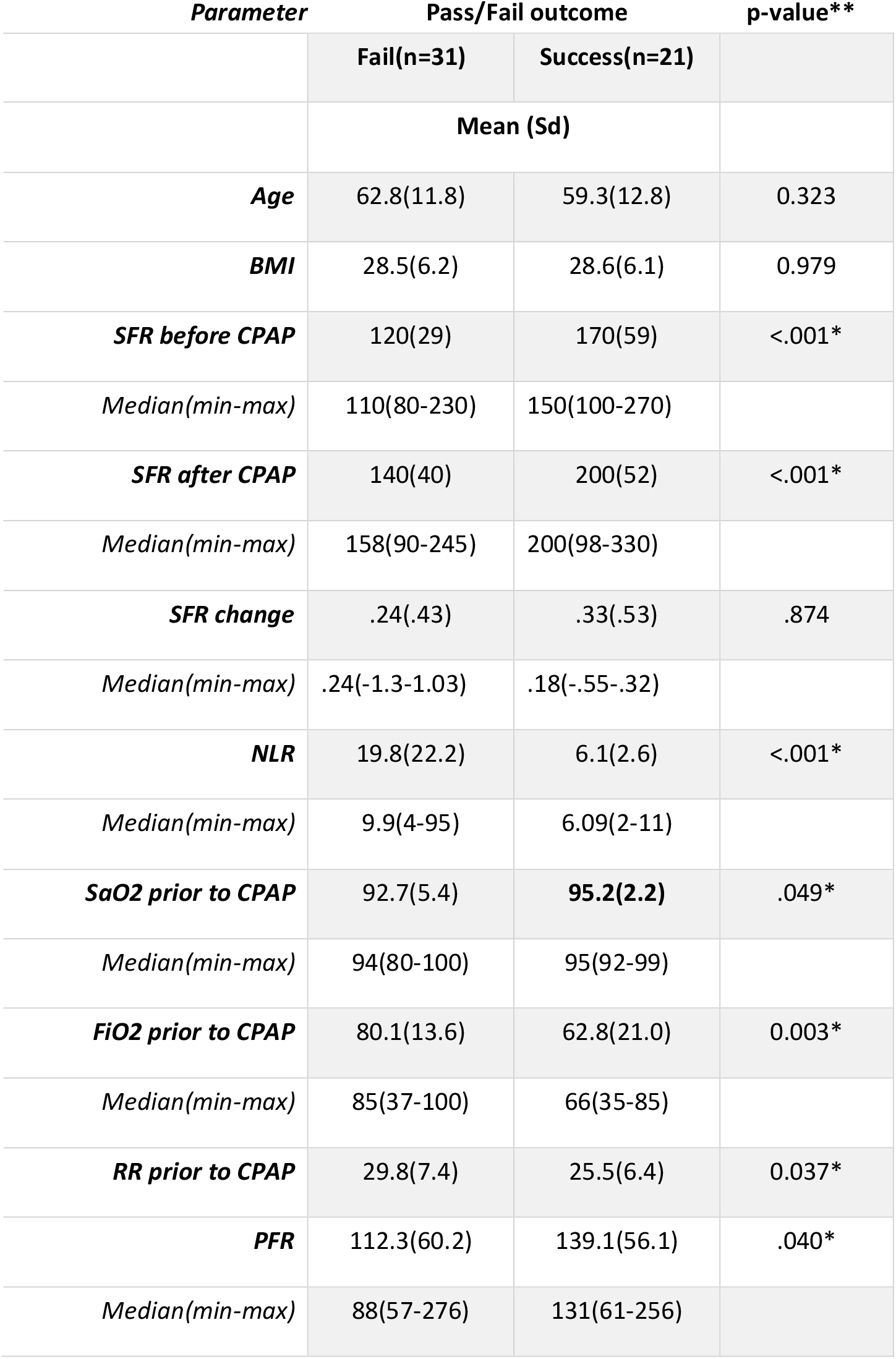
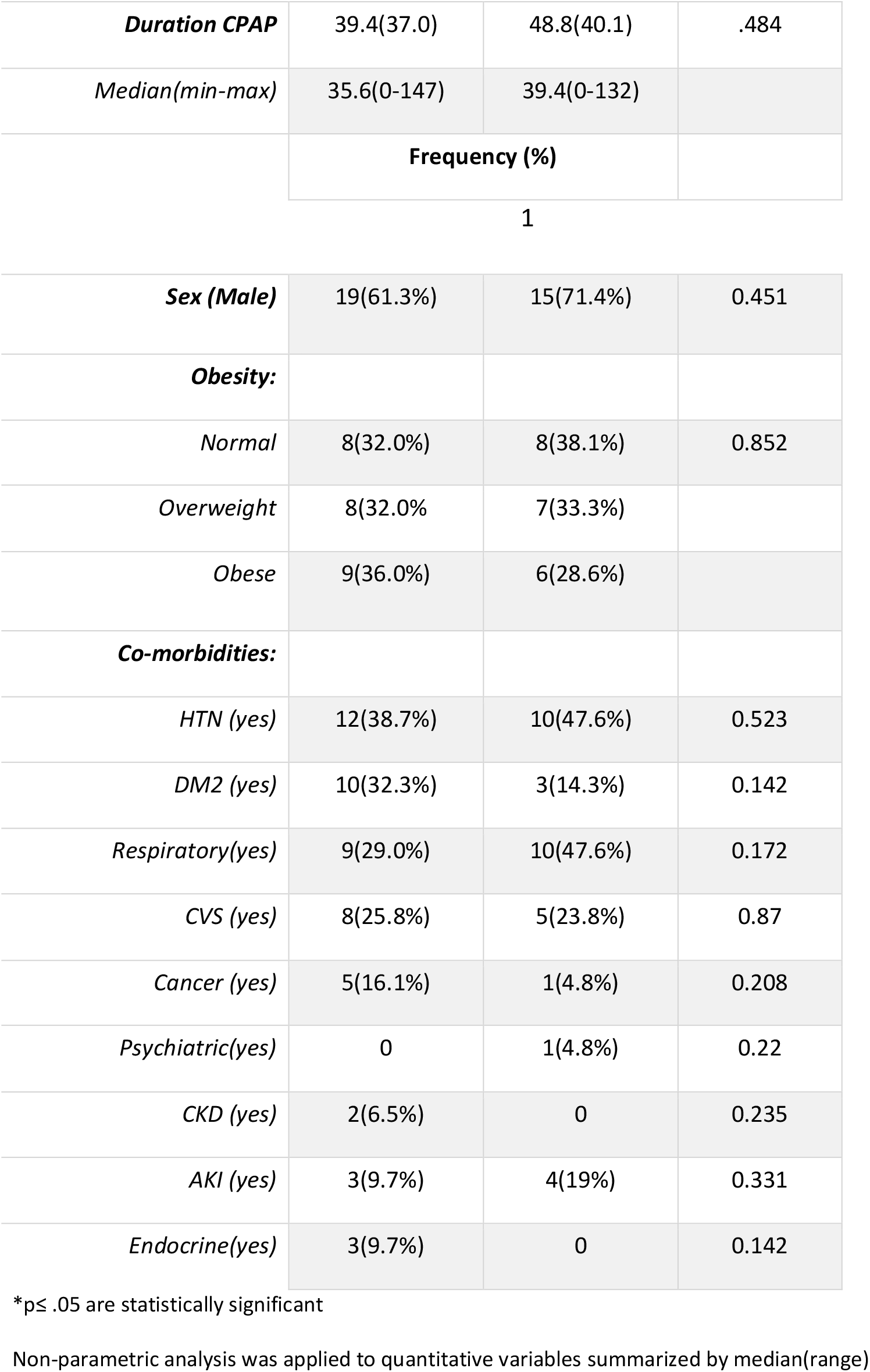
Demographic and clinical characteristics of Patients with COVID19 undergoing CPAP.

Both pre and post CPAP median SFR values were significantly higher in those patients that underwent a successful CPAP trial (p <.05) and median NLR was significantly lower. However, ΔSFR did not differ significantly between both groups (p >.05). For all patients, the mean SFR improved significantly post-CPAP (140±50, p<.001) whether patients failed (p.001) or passed the trial (p .016). We detected also significant positive linear relation between PFR and SFR before (r=.417, p.002) and after CPAP (r =.579, p<.001). NLR was a fair predictor for CPAP success with corresponding sensitivity of 76.2%, specificity of 71% and accuracy of 79% (AUC=0.79, p <.001) (Table 2). A cut-off value less than 8.21 best predicted CPAP success (Figure 2). (OR, 95%CI: 7.38, 1.64-33.19). Moreover, SFR pre-CPAP significantly differentiated success from failure (AUC=0.78, p <0.001). A cut off value of < 114 best predicted CPAP failure. SFR 30-120 minutes post-CPAP was a good predictor of CPAP success (AUC=.807, p<.001) with a sensitivity and specificity of 71.4 and 81.7% respectively. A cut-off value of 180 best distinguished between CPAP success and failure with values ≥ 180 predicting CPAP success. A higher specificity for SFR post-CPAP than sensitivity with large LR+ of 5.53 reveals that it is a good predictor of CPAP success. Comparison of AUC for SFR before and after CPAP revealed no significant difference and both are significantly important predictors for CPAP success (p= .823). PFR is a comparably poor predictor for CPAP success (AUC =.66, p .026) with cut-off value ≥106 predictive of CPAP success (figure 3).

**Table 2:** Predictive performance of different indicators for prognosis of CPAP success outcome in patients with COVID19:

| Parameter | AUC<br>(95% CI) | Optimal Cut-off values | Sig | Sensitivity<br>(%) | Specificity<br>(%) | PPV<br>(%) | NPP<br>(%) | LR+ | LR- |
| --- | --- | --- | --- | --- | --- | --- | --- | --- | --- |
| **NLR** | 0.79<br>(0.67-.91) | ≤ 8.21 | <.001* | 76.2<br>(52.8 -91.8) | 71<br>(52-85.8) | 64<br>(44-86.1) | 81.5<br>(60.6-91.6) | 2.62<br>(1.44-4.48) | 0.33<br>(0.15-.74) |
| **SFR before CPAP** | 0.78<br>(0.66-0.90) | ≥114 | <.001* | 85.7<br>(63.7-97) | 58.1<br>(39.1-75.5) | 58.1<br>(39.1-88) | 85.7<br>(63.7-93) | 2.044<br>(1.30-3.20) | 0.246<br>(0.08-.73) |
| **SFR after CPAP** | .80<br>(.68-.93) | ≥180 | <.001* | 71.4<br>(47.8-88.7) | 87.1<br>(70.2-96.4) | 78.9<br>(56.6-92.2) | 81.8<br>(62.3-94.7) | 5.53<br>(2.13-14.36) | 0.32<br>(0.16-0.65) |
| **PFR ratio** | 0.66<br>(0.51-0.81) | ≥106 | .026* | 76.2<br>(52.8-91.8) | 61.3<br>(42.2-78.2) | 57.1<br>(38.1-82.3) | 79.2<br>(57.1-89.6) | 1.96<br>(1.19-3.25) | 0.38<br>(0.17-.87) |

\*results $p \leq .05$ are statistically significant, 95% CI: 95% confidence interval, PPV: Positive predictive value, NPP: Negative predictive value, LR+: Likelihood ratio for positive test, LR-: Likelihood ratio for negative test.

**Table 3:** Multivariate logistic regression analysis for assessing the contribution of different parameters to CPAP success.

| Parameter | beta | SE | Wald statistic | P value | Adjusted odds ratio (95 %CI) |
| --- | --- | --- | --- | --- | --- |
| **Model1:** |  |  |  |  |  |
| **NLR** <sub>(ref1)</sub> | 2.17 | 0.73 | 8.75 | .006 | 8.75(2.08-36.8) |
| **SFR before CPAP** <sub>(ref2)</sub> | 2.23 | 0.89 | 7.62 | .003 | 9.37(1.9-45.9) |
| **Constant** | -3.02 | 0.86 | 12.3 | <.000 | .049 |
| **Model2:** |  |  |  |  |  |
| **NLR** <sub>(ref1)</sub> | 2.00 | 0.76 | 6.80 | .009 | 7.38(1.64-33.19) |
| **SFR after CPAP** <sub>(ref3)</sub> | 2.55 | 0.79 | 10.39 | .001 | 12.84(2.72-60.62) |
| **Constant** | -2.39 | 0.68 | 12.25 | <.001 | .091 |

Ref1: NLR>8.21; ref2: SFR before CPAP<114; ref3: SFR after CPAP<180
Variable(s) entered on step1, model1: Age, sex, NLR, RR prior to CPAP, PFR and SFR before CPAP.
Variable(s) entered on step1, model2: Age, sex, NLR, RR prior to CPAP, PFR, and SFR after CPAP.
Model1( $\chi^2$ ) =21.07, $p<.001^*$ ; Nagelkerke $R^2$ =45% (model could explain 45% of variance in CPAP success)
Model2( $\chi^2$ ) =24.27, $p<.001^*$ ; Nagelkerke $R^2$ =50.4% (model could explain 50.4% of variance in CPAP success)

**Figure (2):**
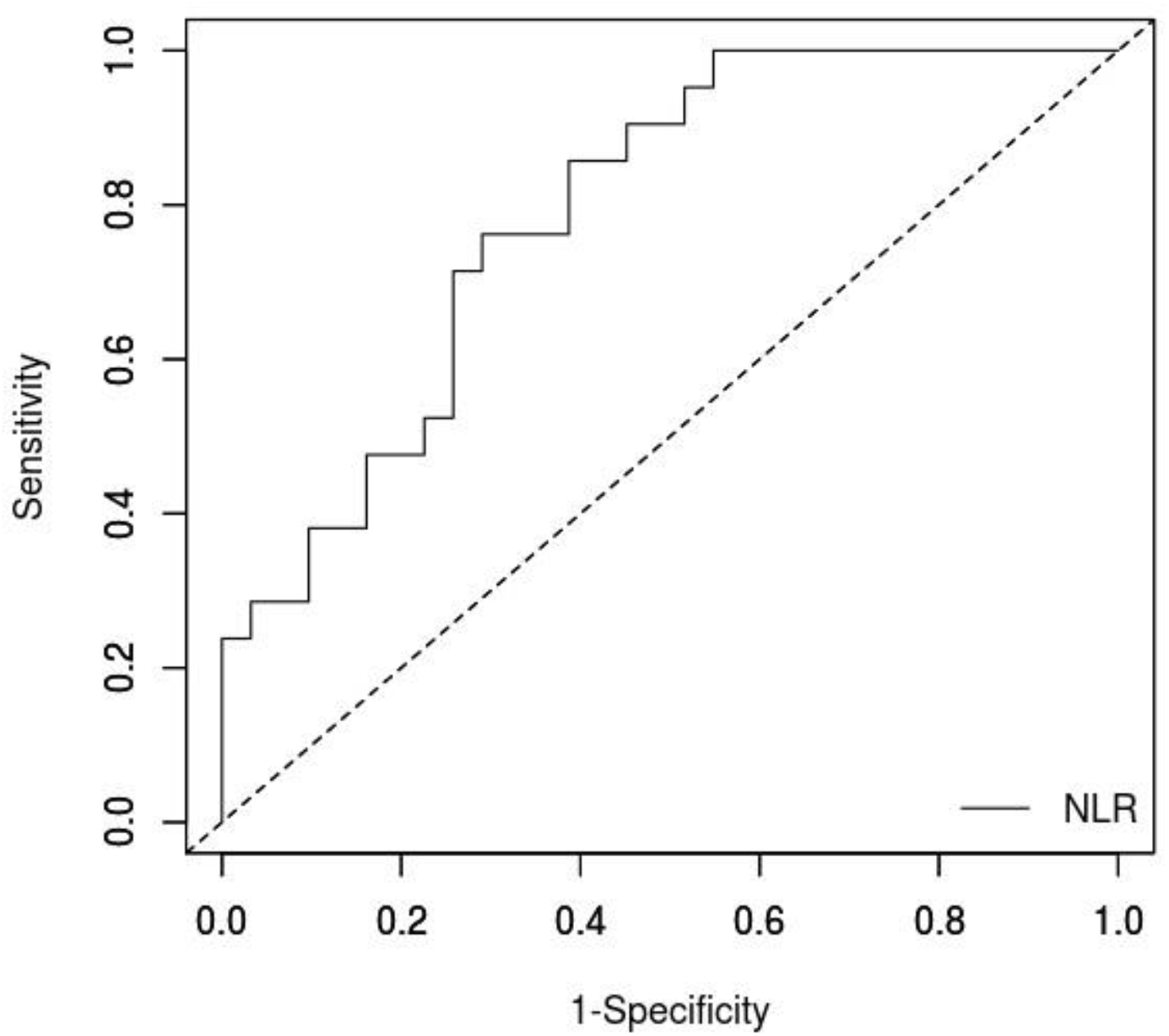
Predictive performance of NLR for CPAP success outcome Small values indicate positive outcome (CPAP success)

**Figure (3):**
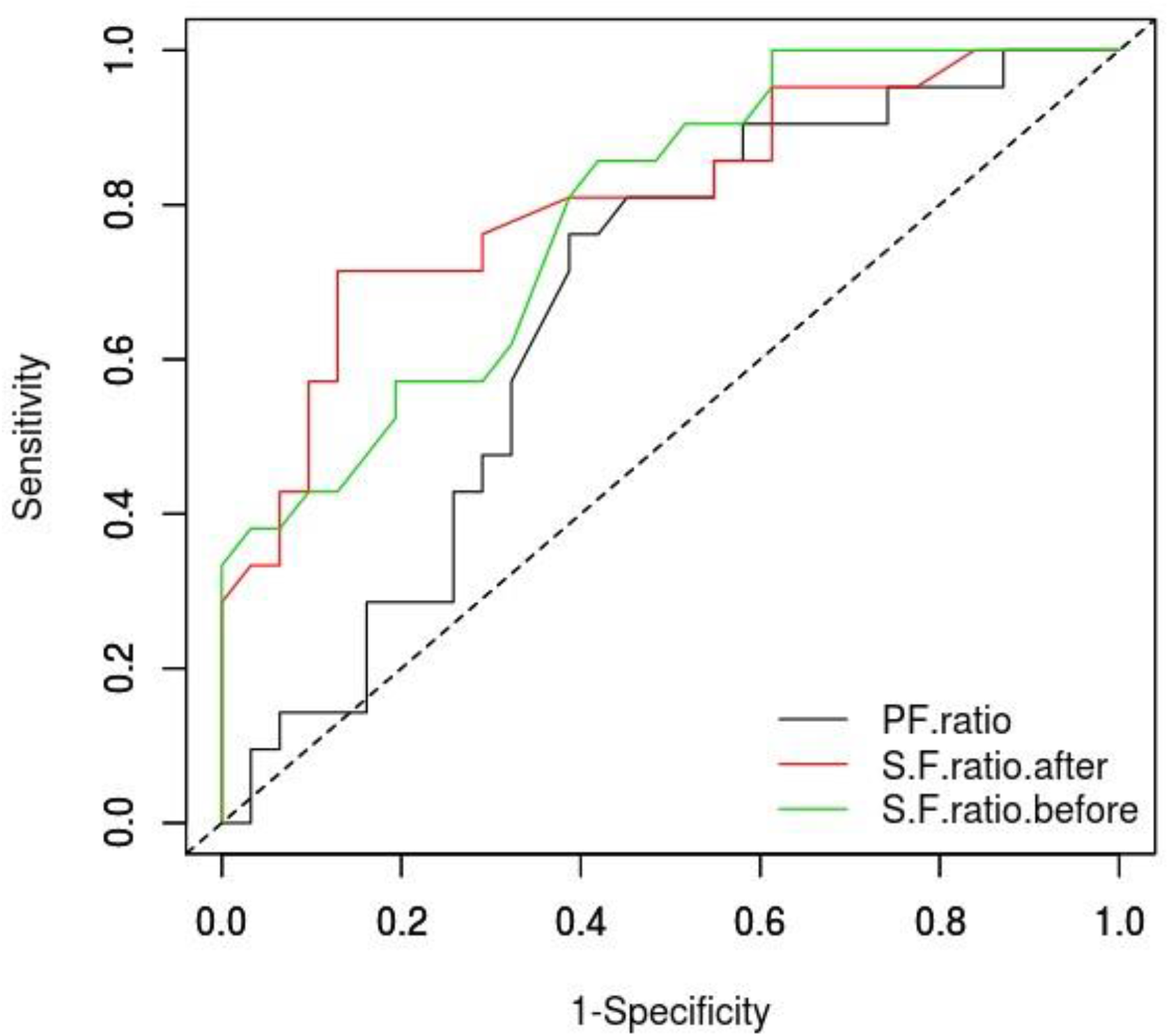
Predictive performance of PFR, SFR before and after CPAP for CPAP success outcome.

95.7% of patients with CPAP success have SFR ≥114 prior to CPAP (p.002) while around 66.7% of CPAP success was achieved in patients with SFR post-CPAP ≥180 (p<.001). 76% of CPAP successes occurred in patients with PFR ≥ 106 (p.008*) and around 76% of the CPAP success group (76.2%) had lower level of NLR ≤ 8.21 (p.001).

Patients with NLR≤ 8.21 had almost 9 times greater chance of CPAP success than those with NLR>8.21 (OR, 95%CI: 8.75, 2.08-36.8). The odds of success are nearly 9 times greater in patients with pre-CPAP SFR ≥114 than patients whose post-CPAP SFR <114 (OR, 95%CI: 9.37, 1.9-45.9). -. SFR 30-120 minutes post-CPAP is the most important predictor of CPAP success (Wald statistic=10.39). The odds of success are about 13 times greater in patients with SFR after CPAP ≥180 than patients whose SFR after CPAP <180 (OR, 95%CI: 12.84, 2.72-60.62).

## Discussion

We report a 40% success of CPAP in COVID-19 patients (n=21/52). We were able to identify 5 significant predictors for CPAP success: RR, PFR, NLR, SFR pre-CPAP; and SFR 30-120 minutes after CPAP (Table 2). An SFR of 180 post-CPAP was the most accurate predictor of CPAP success (AUC = 0.8) (Table 2 & 3, Figure 2 & 3).

Hypoxic COVID-19 patients have a poor prognosis. The current published data on NIPPV in COVID-19 patients is scarce and mainly reports survival as an endpoint. Xie et al reported that an SpO_2_ less than 90% despite O_2_ therapy is associated with high mortality. A quarter of such patients required ventilatory support including IMV for 12% of them. (14) They reported 47% NIPPV success rate (7 out of 13). (14) Chen et al showed that just 26 out of 102 patients after NIPPV survived. (15) Yang et al showed that among 52 ITU patients, 29 were managed by NIPPV and only 6 (20%) survived. (16) Our outcomes were better than previously reported.

The overall survival rate after CPAP (whether subsequently intubated or not) was 80% in those patients that were for full escalation (33 out of 41). Survival rate across the whole cohort irrespective of ceiling of care was 65% (34 out of 52). In our group of patients, among the 41 patients who had a decision “For Full Escalation”, 20 patients passed their CPAP trial successfully avoiding intubation and ITU admission (48.8%). Ten patients failed NIPPV and were palliated (decision not to intubate).

Applying CPAP can improve oxygenation, recruit collapsed lung regions and ameliorate the work of breathing. The latter can protect from patients’ self-inflicted lung injury (P-SILI) and diaphragmatic injury. A recent study showed that 64% of COVID-19 patients have recruitable lungs on day 1 after ITU admission which may explain the benefit of CPAP. (17) An additive benefit may be observed in patients with concomitant heart failure (reduced preload). A recent meta-analysis showed that NIPPV can decrease both the intubation rate and mortality for patients with PFR of 200-300 mmHg, and when using helmets. (14) Another observed benefit was the reduction of ITU and ventilator days which is of particular importance when there is a severe strain on resources. (18) Most significantly, a recent systematic review based on current evidence suggests that the use of NIPPV in AHRF, similar to IMV, likely reduces the risk of mortality but may increase the risk of transmission of COVID-19 to healthcare workers. (19) The avoidance of endotracheal intubation carries many benefits not least of which is the reduced burden on scarce ITU resources during a pandemic.

Our data suggest that monitoring should focus mainly on 2 parameters: (1) Work of breathing and (2) Oxygenation. CPAP helps recruiting more functioning lung units. (17) Patients with good lung compliance can generate high tidal and minute volume before feeling short of breath and the RR started to increase significantly. In our study, we showed a significant difference in the RR existed between the failure and success groups before CPAP (Mean±SD 29.8±7.4 versus 25.5± 6.4 cycles per minute respectively, p=0.037) (Table 1).

We have demonstrated the value of SFR. Monitored before and 30-120 minutes after the CPAP trial, it showed high sensitivity and specificity in detecting CPAP outcome. In our opinion, such an approach can significantly improve the safety of the CPAP trial out of the ITU settings. SFR offers many advantages over PFR. It is based on a non-invasive, continuous and real time pulse oximeter monitoring. By using CPAP and monitoring SFR, there is reduced need of ABG, cost and complications related to this procedure. Moreover, SFR is non-invasive measurement, easily monitored remotely thus reducing exposure to staff from risk of infection related to aerosol generating procedures (AGPs). However, we appreciate many limits of the SFR secondary to the potential inaccuracy of the pulse oximeter in certain conditions: motion, low perfusion, dyshaemoglobinaemias, clubbing and nail polish. (20) The accuracy of SFR parallels that of the SpO_2_: less accurate and less reflective the PaO_2_ when the SpO_2_ saturation becomes less than 90%. (21) We recommend that the SFR should not be relied on solely in patient with SpO_2_ less than 90%. A constant relation between SpO_2_ and PaO_2_ when in the range between 90 and 100% had been suggested and fits our algorithm and patients. (21) The target SpO_2_ was 92 to 96% for all our patients according to British Thoracic Society (BTS) guidelines. (22)

The SFR has previously been shown to have a promising role to predict ITU admission. (23) Rice et al were able to describe a relation between PFR and SFR. An SFR of 235 and 315 corresponded to PFR of 200 and 300 with a sensitivity of 85% and 91%; and specificity of 85% and 91% respectively. (24) Bilan et al also were able to demonstrate the reliability of SFR for the diagnosis of moderate ARDS, in substitute of the PFR. They demonstrate that an SFR of 181 and 235, predict a PFR of 200 and 300, respectively. (25) Using equations suggested in the previous 2 studies, we can suggest that our post-CPAP SFR cut-off of 180, corresponds to PFR of 200 mmHg and moderate ARDS.

Lastly, our results show NLR as a strong predictor of CPAP success. This is related to its significance in predicting disease severity. Liang et al demonstrated that NLR was one of the 10 independent and statistically significant predictors of critical illness (OR, 1.06; 95% CI, 1.02-1.10; *P* = .003) in COVID-19 patients. (26) Liu et al and Yang et al also showed that NLR is a significant risk factor for poor outcome. (27,28) A meta-analysis of 5 studies showed NLR to be associated with COVID-19 severity. (29) The NLR in COVID-19 reflects inflammation which stimulates neutrophilia from one side and lymphocyte apoptosis from the other side. (30)

Based on our results, we suggested a modified CPAP management protocol in COVID-19 patients. (Fig 4) The modified algorithm needs further confirmation in a larger study.

**Figure (4):**
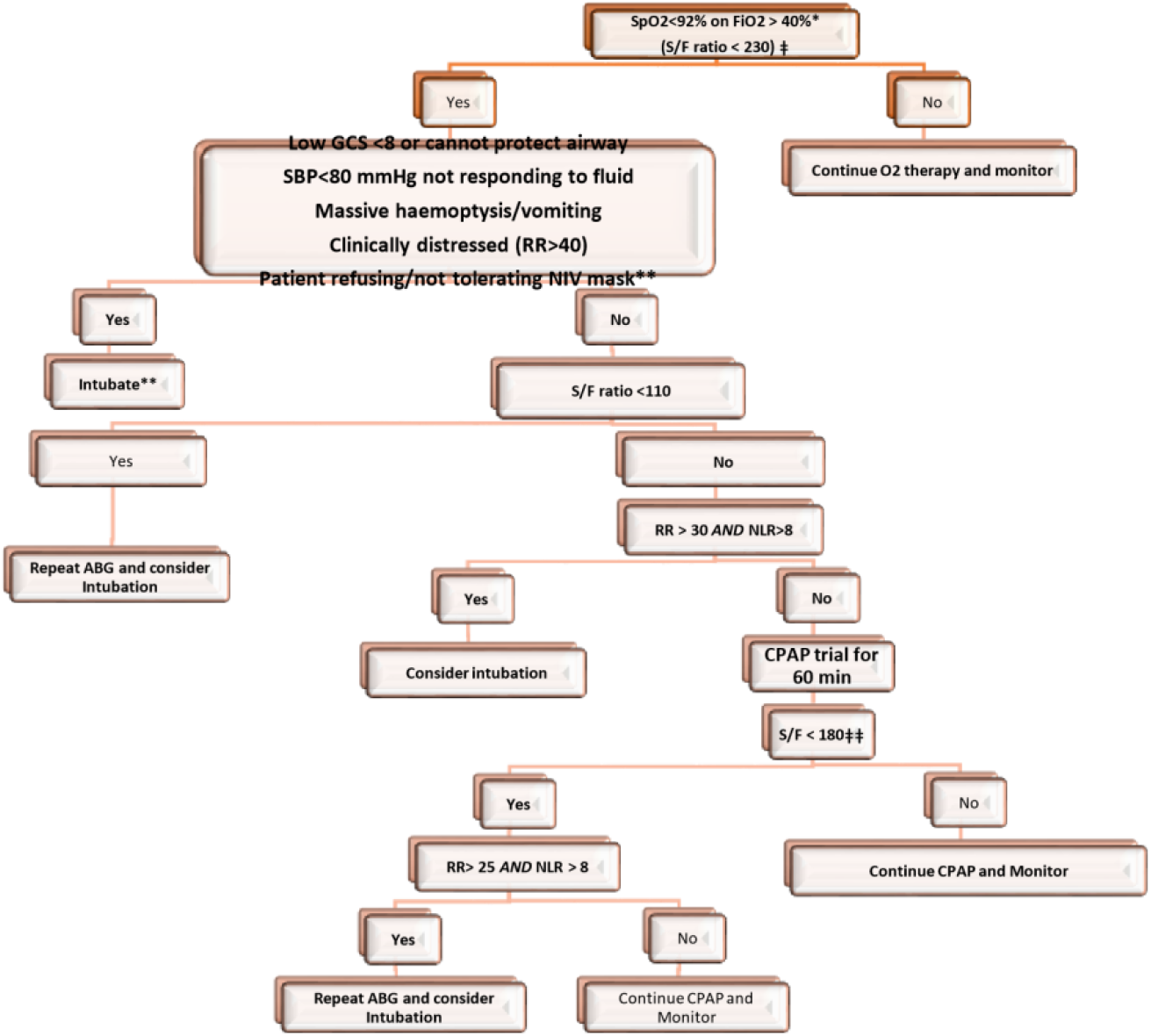
Decision tree clinical algorithm for prediction of CPAP Success. *Target SpO2 88-92% if risk of T2RF ** If no ceiling of care ǂ First Decision tree for prediction of CPAP success before CPAP procedure, accuracy = 78.8% and corresponding AUC= 0.870 ǂǂ Second Decision tree for prediction of CPAP success after CPAP procedure, accuracy = 87.7% and corresponding AUC= 0.867

### Decision tree algorithm for prediction of CPAP Success

Before a CPAP trial, the best predictor of success is SFR. If a patient has SFR ≥230, probable outcome is CPAP success but if SFR <110, patients are more likely to fail on CPAP. Patients with SFR between 110-230 should have RR prior to CPAP measured and patients are more likely to succeed on CPAP if RR≥ 30 per minute.

After CPAP trial, patients are monitored and if SFR is above 180, probable outcome is CPAP success. If SFR ≤ 180, then the next step is to check NLR and if it is >8.21, probable outcome is CPAP failure. However, patients with NLR ≤8.21 with RR ≥ 25/ min are more likely to pass on CPAP trial. Patients with NLR ≤8.21 with RR < 25/ min are predicted to fail on CPAP trial. (Figure 4)

### Limits

Our study is not without limitations. First, it is a retrospective observational study conducted on a small group of patients. Our results need confirmation in a larger prospective work. Second, many mechanisms can contribute to AHRF even in a single disease (e.g. Pneumonia, ARDS, but also pulmonary embolism which is not rare in COVID-19). Third, it remains to be determined if the CPAP trial affected the outcome of intubated patients in comparison to those promptly intubated from the start. Furthermore, we did not use High Flow Nasal Cannula (HFNC) which may have a role. A randomised controlled trial may be needed for full comparison between the 3 treatment modalities of HFNC, CPAP and early IMV.

### Conclusion

CPAP has a significant role in the management of COVID-19 patients presenting with AHRF. We have demonstrated that an experienced medical team following protocol driven management can successfully use CPAP to manage patients with COVID-19 and AHRF. In our group of patients, its success rate was 40%. SFR, PFR, NLR and RR are predictors of such success. SFR is a non-invasive, real-time measurement which can be used effectively to monitor these patients before and after CPAP to identify likelihood of success and reduce the need for frequent ABGs.

The proposed modified CPAP management algorithm provides an initial step for better identifying those patients who will likely succeed on CPAP therapy. Thus, reducing the need for unnecessary IMV, and its associated complications, which can relieve pressure on scarce ITU resources during a pandemic. This algorithm will strengthen decision making should we encounter a second wave of COVID-19.

## Data Availability

All data available under reasonable request

